# Effectiveness of BNT162b2 (Comirnaty, Pfizer-BioNTech) COVID-19 booster vaccine against covid-19 related symptoms in England: test negative case-control study

**DOI:** 10.1101/2021.11.15.21266341

**Authors:** Nick Andrews, Julia Stowe, Freja Kirsebom, Charlotte Gower, Mary Ramsay, Jamie Lopez Bernal

**Author notes:** Joint first authors (Nick Andrews, Julia Stowe).

## Abstract

**Background:** In September 2021, the UK Government introduced a booster programme targeting individuals over 50 and those in a clinical risk group. Individuals were offered either a full dose of the BNT162b2 (Comirnaty, Pfizer-BioNTech) vaccine or a half dose of the mRNA-1273 (Spikevax, Moderna) vaccine, irrespective of the vaccine received as the primary course

**Methods:** We used a test-negative case-control design to estimate the Vaccine Effectiveness (VE) of the booster dose BNT162b2 (Comirnaty, Pfizer-BioNTech) in those aged over 50 against symptomatic disease in post booster time intervals compared to individuals at least 140 days post a second dose with no booster dose recorded. In a secondary analysis, we also compared to unvaccinated individuals and to the 2 to 6 day period after a booster dose was received. Analyses were stratified by which primary doses had been received and any mixed primary courses were excluded.

**Results:** The relative VE estimate in the 14 days after the BNT162b2 (Comirnaty, Pfizer-BioNTech) booster dose, compared to individuals that received a two-dose primary course, was 87.4 (95% confidence interval 84.9-89.4) in those individuals who received two doses ChAdOx1-S (Vaxzevria, AstraZeneca) as a primary course and 84.4 (95% confidence interval 82.8-85.8) in those individuals who received two doses of BNT162b2 (Comirnaty, Pfizer-BioNTech) as a primary course. Using the 2-6 day period post the booster dose as the baseline gave similar results. The absolute VE from 14 days after the booster, using the unvaccinated baseline, was 93.1(95% confidence interval 91.7-94.3) in those with ChAdOx1-S (Vaxzevria, AstraZeneca) as their primary course and 94.0 (93.4-94.6) for BNT162b2 (Comirnaty, Pfizer-BioNTech) as their primary course.

**Conclusions:** Our study provides real world evidence of significant increased protection from the booster vaccine dose against symptomatic disease in those aged over 50 year of age irrespective of which primary course was received.

## Background

Real world effectiveness data demonstrated high levels of short-term protection by COVID-19 vaccines against clinical disease and, more so, against severe outcomes including hospitalization and death (1-7). Nevertheless, there is now evidence that protection against symptomatic disease wanes over time (8, 9). Booster doses have now been implemented in the UK in order to combat the rise in COVID-19 cases and the additional threat of the winter 2021 influenza season.

We recently reported that vaccine effectiveness against symptomatic disease peaked in the early weeks after the second dose and then fell to 47.3 (95% CI 45 to 49.6) and 69.7 (95% CI 68.7 to 70.5) by 20+ weeks against the Delta variant for ChAdOx1-S (Vaxzevria, AstraZeneca) and Pfizer-BioNTech (BNT162b2/ Comirnaty®), respectively. Vaccine effectiveness against severe disease outcomes remained high to 20+ weeks after vaccination in most groups, nevertheless, greater waning was seen in older adults and those with underlying medical conditions compared to young, healthy adults (8).

In the UK, COVID-19 booster vaccines were introduced on 14 September 2021. Using evidence from the COV-BOOST trial, which demonstrated that the mRNA vaccines provide a strong booster effect with low reactogenicity, regardless of the vaccine given in the primary course (10), the UK Joint Committee on Vaccination and Immunisation (JCVI) recommended either a BNT162b2 or a half dose (50µg) of mRNA-1273 (Spikevax, Moderna) vaccine to be given as a booster dose no earlier than 6 months after completion of the primary vaccine course (10). In this initial phase of the UK booster programme the following groups were eligible: all adults over 50 and those 16-49 years with underlying health conditions that put them at higher risk of severe COVID-19, adult carers and adult household contacts (aged 16 or over) of immunosuppressed individuals, and healthcare workers.

In this study, we aimed to estimate the effectiveness of booster vaccination against symptomatic disease in adults aged 50 years and older.

## Methods

### Study Design

We used a test-negative case-control design to estimate vaccine effectiveness of a booster dose of BNT162b2 vaccine against PCR-confirmed symptomatic disease. We compared vaccination status in symptomatic adults over 50 years of age with PCR-confirmed SARS-COV-2 infection with the vaccination status in individuals which reported symptoms but had a negative SARS-COV-2 PCR test. As mRNA-1273 vaccine, as a primary course, was not made available until later in the vaccine programme insufficient time had elapsed for a booster dose to be indicated in this group In addition, there were very few individuals that had received the half dose (50µg) of mRNA-1273 vaccine as a booster dose so we were unable to assess the VE of this vaccine in our study.

### Data Sources

#### Vaccination data

The National Immunisation Management System (NIMS) (11) contains some demographic information on the whole population of England who are registered with a GP in England and is used to record all COVID-19 vaccinations. These data were accessed on 01 November 2021. The information used from NIMS was all dates of COVID-19 vaccination, vaccine manufacturer for each dose. Demographic data such as sex, date of birth, ethnicity, and residential address was extracted. Addresses were used to determine index of multiple deprivation quintile and were also linked to Care Quality Commission registered care homes using the unique property reference number. NIMS also contained data on geography (NHS region), risk groups status, clinically extremely vulnerable, and health/social care worker.

Booster doses were identified as being a third dose 140 days or more after a second dose and given after 13^th^ September 2021. Individuals with four or more doses of vaccine, a mix of vaccines in their primary schedule or less than 19 days between their first and second dose were excluded.

#### COVID-19 testing data

SARS-CoV-2 Testing Polymerase-chain-reaction (PCR) testing for SARS CoV-2 in the United Kingdom is undertaken by hospital and public health laboratories, as well as by community testing with the use of drive through or at-home testing, which is available to anyone with symptoms consistent with Covid-19, is a contact of a confirmed case, for care home staff and residents or who has self-tested as positive using a lateral flow device. Initially data on all positive and negative tests for the period 08 December 2020 to 29 October 2021 were extracted for individuals aged ≥ 50 years on 31 August 2021. Any negative tests taken within 7 days of a previous negative test, or where symptoms were recorded, with symptoms within 10 days of symptoms for a previous negative test were dropped as these likely represent the same episode. Negative tests taken within 21 days before a positive test were also excluded as these are likely to be false negatives. Positive and negative tests within 90 days of a previous positive test were also excluded. Participants contributed a maximum of four randomly chosen negative test results in the follow-up period. Data were restricted to persons who had reported symptoms and gave an onset date. Only persons who had undergone testing within 10 days after symptom onset were included in order to account for reduced sensitivity of PCR testing beyond this period. A small number of positive samples where sequencing was done and they were found not to be the Delta variant were excluded. Finally, only samples taken from 13 September 2021 (week 37, 2021) were retained for analysis.

#### Linkage of testing data to NIMS

Testing data were linked to NIMS on 01 November 2021 using combinations of National Health Service number (a unique identifier for each person receiving medical care in the United Kingdom), date of birth, surname, first name, and postcode using deterministic linkage with >95.5% uniqueness. The NIMS denominator file included information on potential confounding variables related to targeted populations.

### Statistical analysis

Analysis was by logistic regression with the PCR test result as the dependent variable where those testing positive are cases and those testing negative controls. Vaccination status was included as an independent variable and effectiveness defined as 1-odds of vaccination in cases/odds of vaccination in controls.

Vaccine effectiveness was adjusted in logistic regression models for age (5 year bands), sex, index of multiple deprivation (quintile), ethnic group, care home residence status, geographic region (nhs region), period (calendar week of onset), health and social care worker status, clinical risk group status, clinically extremely vulnerable, severely immunosuppressed, and previously testing positive. These factors were all considered potential confounders so were included in all models.

Analyses were stratified by which primary doses had been received, ChAdOx1-S or BNT162b2 and any mixed primary courses were excluded. Vaccine effectiveness was assessed for each primary course of vaccine with a BNT162b2 booster in 0-1, 2-6, 7-13, 14+ day post booster vaccine intervals. In the primary analysis, those that had received the booster were compared to individuals who had received two primary doses with at least 140 days prior to the onset but with no booster dose recorded. In secondary analyses, we also compare to completely unvaccinated individuals and to the 2-6 day period after the booster was received. The 2-6 day period was selected after plotting the data on case and control numbers after the booster dose and to avoid days 0 and 1 post booster when vaccine reactogenicity may affect the case-control ratio (figure 1). The analyses comparing to two doses or the 2-6 day post booster period measures relative effectiveness to two doses, whilst the comparison to unvaccinated is absolute effectiveness of two doses and a booster. In the analysis comparing to unvaccinated we also assessed the remaining effectiveness of two doses at least 140 days (20 weeks) post second dose.

**Figure 1:**
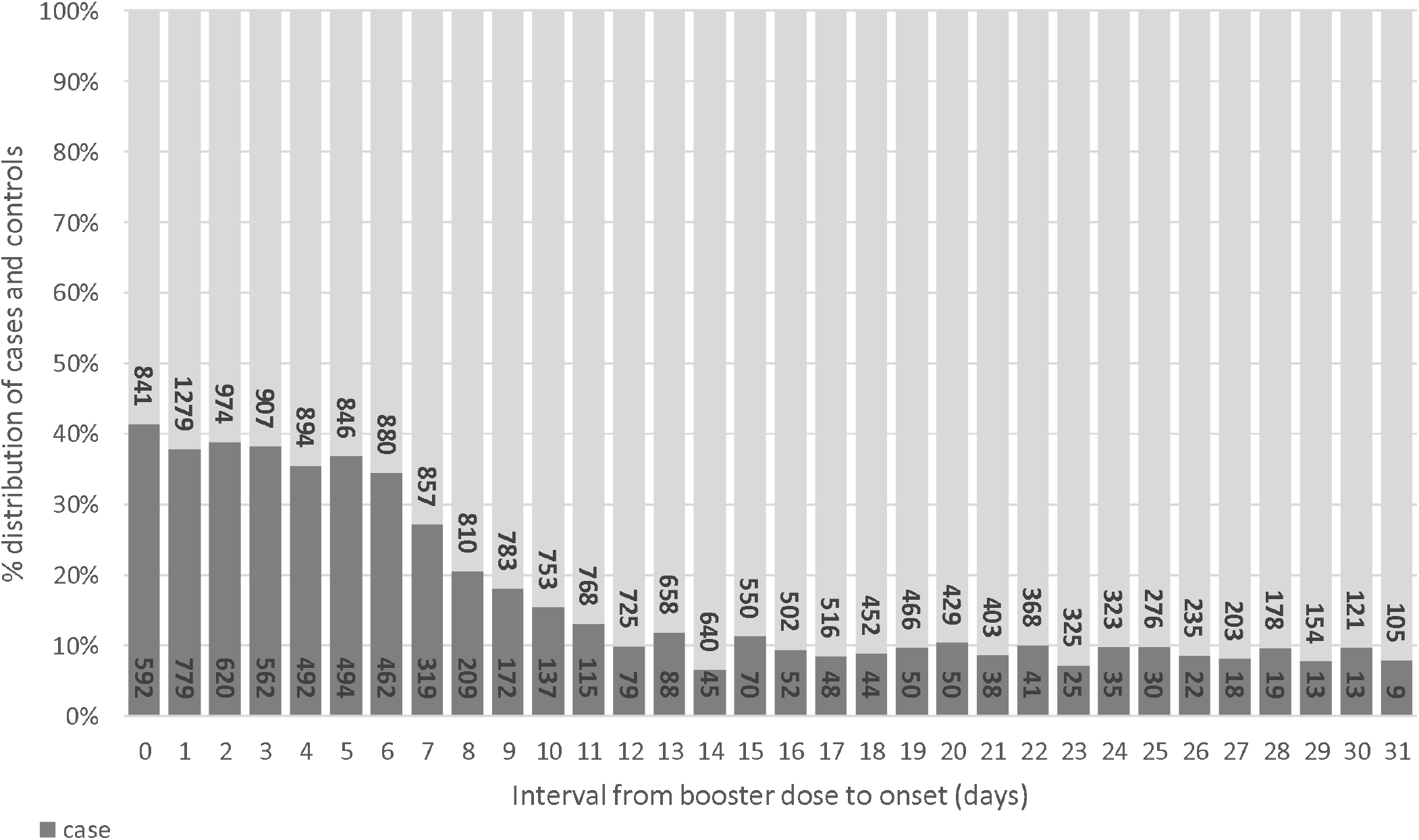
Cases and controls by interval from booster to onset.

## Results

### Descriptive Statistics and characteristics

From week 37 onwards there were a total of 271,747 eligible tests in those aged 50 years and over, with a test date within 10 days of their symptom onset date and had linked to the National Immunisation Management system, with a 95.7% match rate. Of these 13,569 (5.0%) were unvaccinated, 149,434 received ChAdOx1-S 140 days post a second dose, 84,506 received BNT162b2 140 days post a second dose. Of those that had received a booster dose BNT162b2 6,716 had received an ChAdOx1-S primary course and 17,521 received a BNT162b2 primary course. A description of the test positive and negative cases is given in Table 1.

**Table 1:**
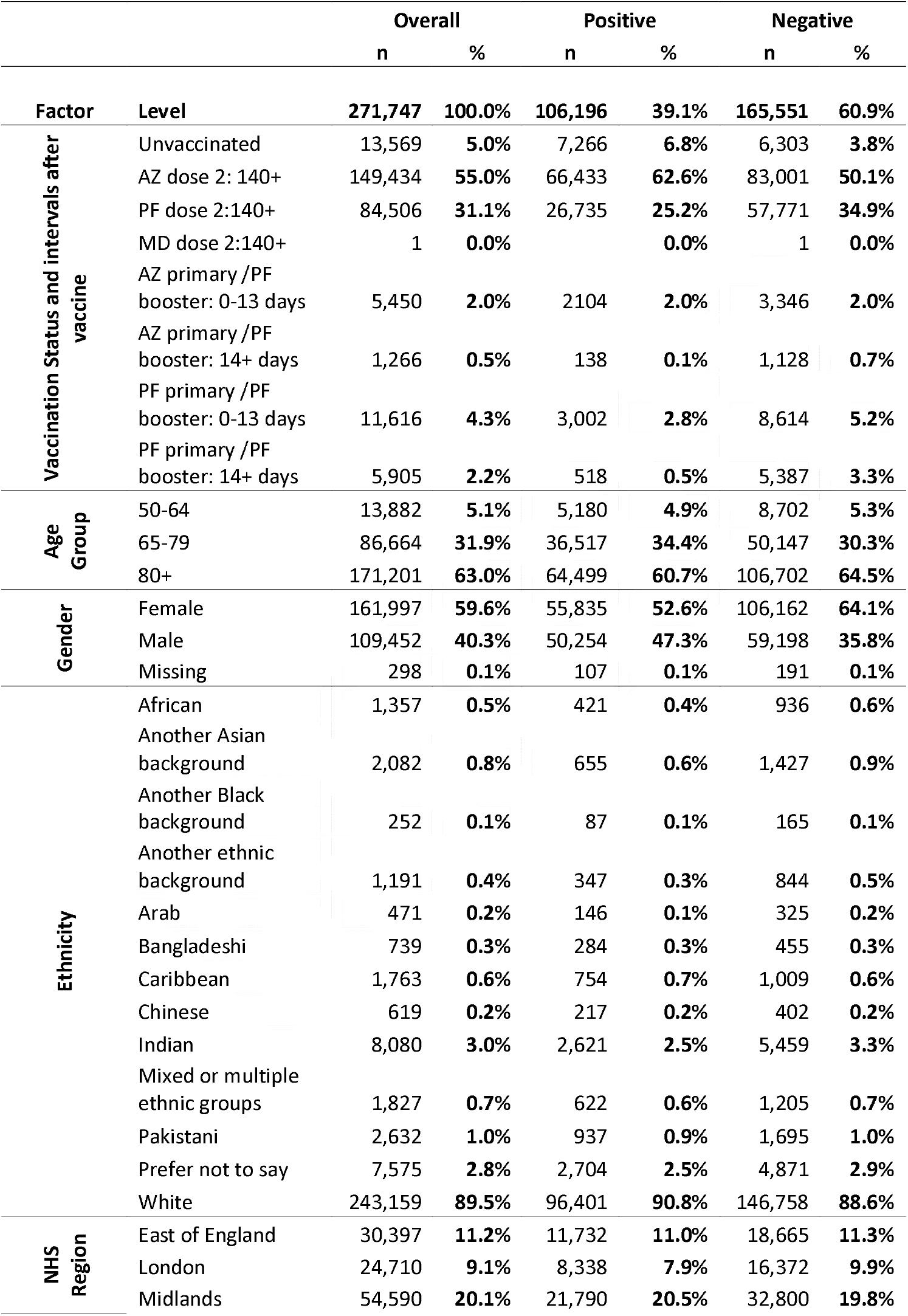
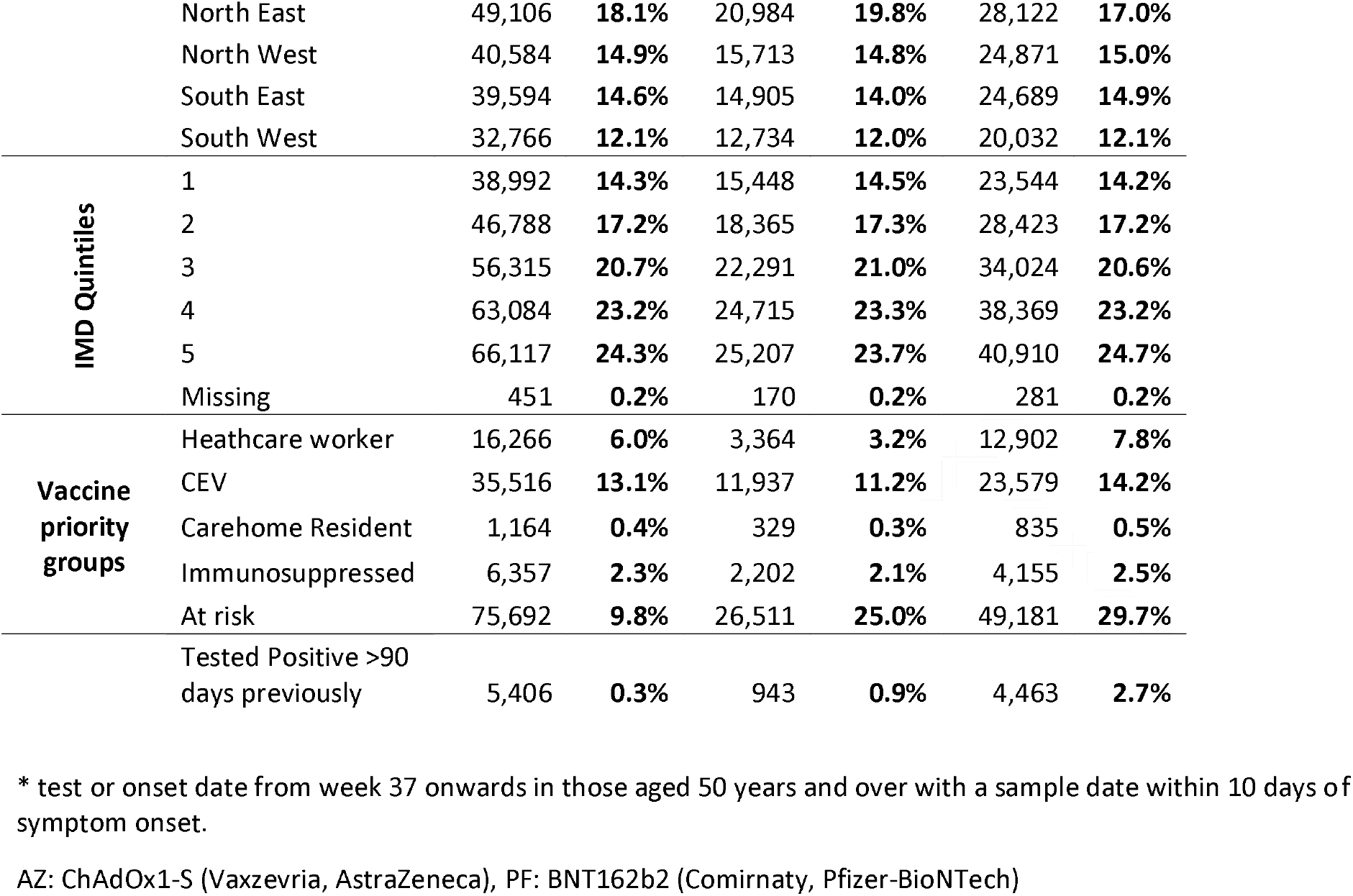
Descriptive characteristics of positive and negative test results in individuals tested for SARS-CoV-2 in Engla the study population. *.

### Vaccine effectiveness estimates

An overall effect on the proportion of cases and controls can be seen from around day 7 after the booster dose and stabilises at day 11 (figure 1). Vaccine effectiveness of a BNT162b2 booster dose relative to those that had received only two doses was 87.4% (95% confidence interval 84.9-89.4) where the primary course was ChAdOx1-S and 84.4% (95% confidence interval 82.8-85.8) where BNT162b2 was used as the primary course (table 2 & figure 2).

**Table 2:** Vaccine effectiveness against symptomatic disease for the BNT162b2 (Comirnaty, Pfizer-BioNTech) booster vaccine in England. Table values are VE (95% CI).

| Primary Course (with second dose 140+ days before) | Interval since PF Booster | Controls | Cases | rVE (140+ days post dose 2 baseline) | rVE (dose 3: 2-6 days post booster baseline) | VE (unvaccinated baseline) |
| --- | --- | --- | --- | --- | --- | --- |
| Unvaccinated | No booster | 6303 | 7266 |  |  | baseline |
| 2AZ | No booster | 83001 | 66433 | Baseline |  | 44.1 (41.9 to 46.1) |
| 2AZ | 0-1 days | 694 | 611 | -0.3 (-12.3 to 10.4) |  | 44.9 (38 to 51) |
| 2AZ | 2-6 days | 1328 | 1095 | 12.9 (5.3 to 19.9) | baseline | 52.4 (47.9 to 56.5) |
| 2AZ | 7-13 days | 1324 | 398 | 68.3 (64.4 to 71.7) | 63.6 (58.1 to 68.3) | 82.8 (80.6 to 84.7) |
| 2AZ | 14+ days | 1128 | 138 | 87.4 (84.9 to 89.4) | 85.5 (82.4 to 88.1) | 93.1 (91.7 to 94.3) |
| 2PF | No booster | 57771 | 26735 | Baseline |  | 62.5 (61.0 to 63.9) |
| 2PF | 0-1 days | 1422 | 758 | -7.1 (-17.5 to 2.4) |  | 59.5 (55.3 to 63.3) |
| 2PF | 2-6 days | 3167 | 1525 | 9.9 (3.8 to 15.7) | baseline | 65.8 (63.2 to 68.2) |
| 2PF | 7-13 days | 4025 | 719 | 68.3 (65.5 to 70.8) | 64.8 (61 to 68.2) | 87.9 (86.7 to 88.9) |
| 2PF | 14+ days | 5387 | 518 | 84.4 (82.8 to 85.8) | 82.6 (80.6 to 84.5) | 94.0 (93.4 to 94.6) |

AZ: ChAdOx1-S (Vaxzevria, AstraZeneca), PF: BNT162b2 (Comirnaty, Pfizer-BioNTech), VE: vaccine effectiveness compared to zero doses, rVE: relative vaccine effectiveness compared to dose 2 (either 140+ days post dose 2 with no booster or 140+ days post dose 2 and 2-6 days after he booster).

**Figure 2:**
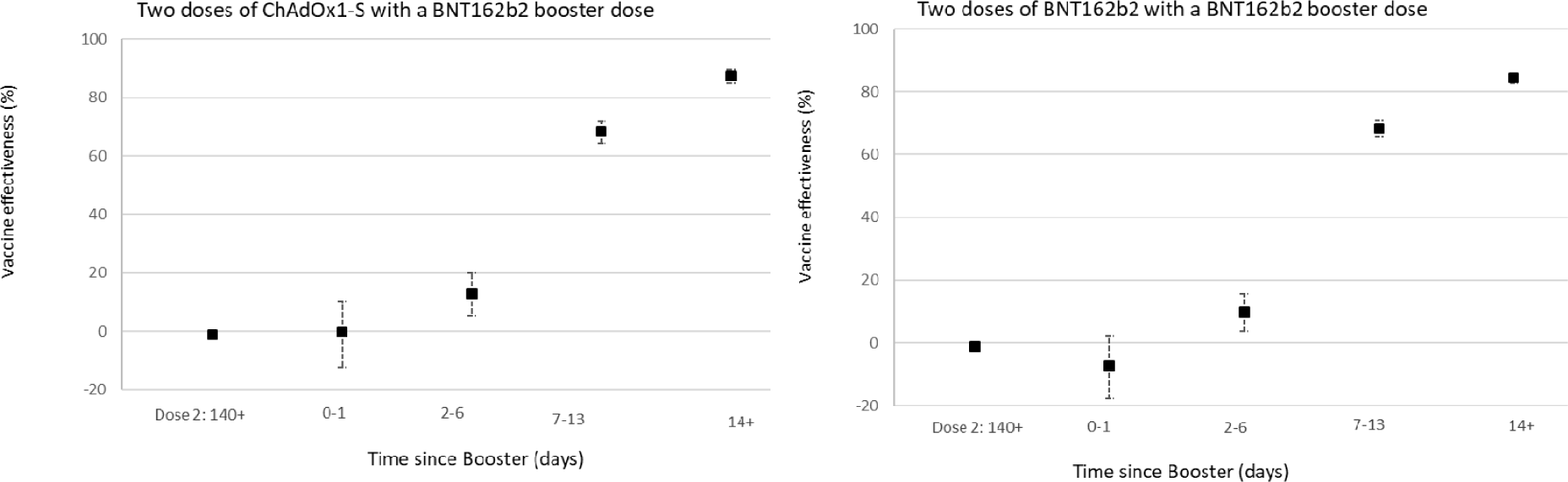
Relative vaccine effectiveness estimates in time intervals post booster according to primary course: 140+ days post dose 2 as baseline (set at 0% VE)

In the secondary analysis, which used the 2-6 day period post the booster dose as the baseline similar results were reported to the primary analysis with a relative VE from 14 days after the booster dose of 85.5% (95% confidence interval 82.4 -88.1) for ChAdOx1-S and 82.6% (95% confidence interval 80.6-84.5) for BNT162b2 as the primary course (table 2 & figure 3). In the analysis using the unvaccinated individuals as the baseline, the booster dose was associated with an absolute VE from 14 days after vaccination to 93.1% (95% confidence interval 91.7-94.3) after an ChAdOx1-S primary course and 94.0 (95% confidence interval 93.4-94.6) after a BNT162b2 primary course (table 2 & figure 4). In the analysis using the unvaccinated baseline the effectiveness of two doses of ChAdOx1-S and BNT162b2 ≥20 weeks after being given were 44.1% and 62.5%, respectively (table 2 & figure 4).

**Figure 3:**
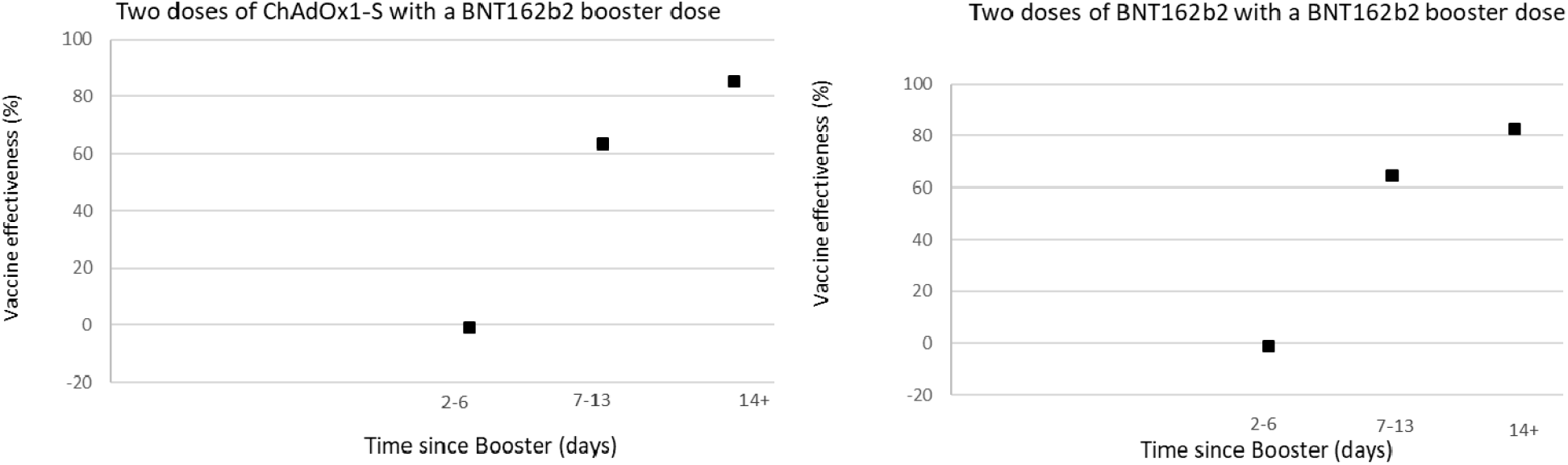
Relative vaccine effectiveness estimates in time intervals post booster according to primary course: 2-6 days post booster as baseline (set at 0% VE)

**Figure 4:**
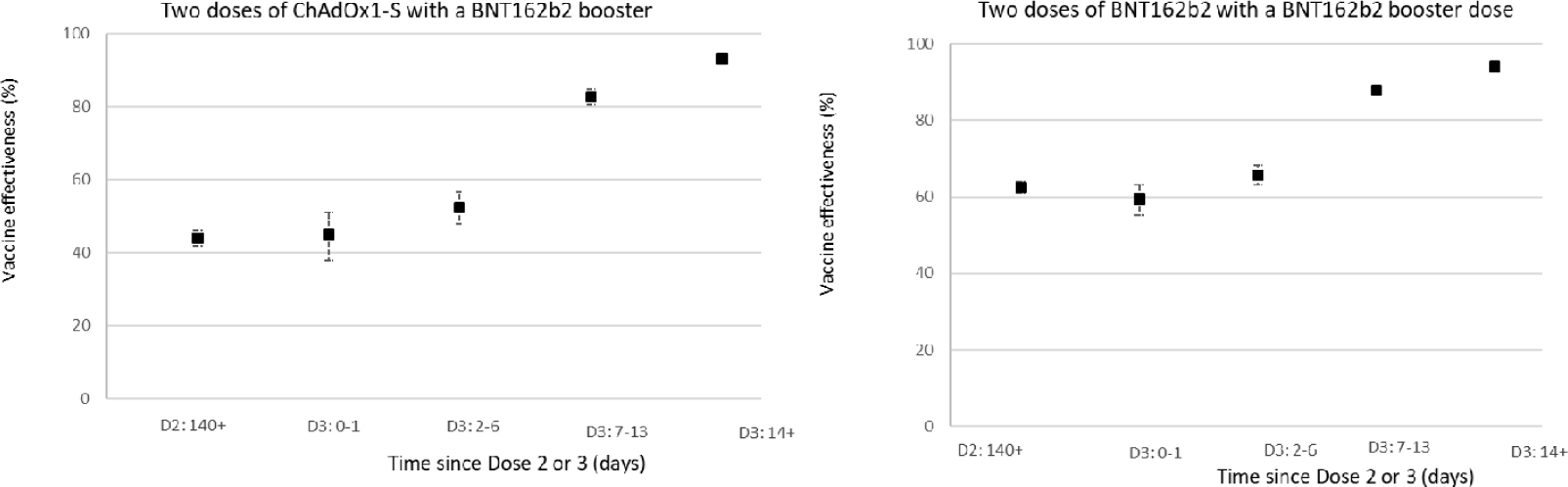
Vaccine Effectiveness estimates for at least 140 days post dose 2 (given with no booster) or for time intervals post dose 3 (booster) according to primary course: Unvaccinated as baseline.

## Discussion

### Key findings

This study provides evidence of a significant increase in protection against symptomatic COVID-19 with a booster dose of BNT162b2 following a primary course of either BNT162b2 or ChAdOx1-S in adults aged 50 years and older. Vaccine effectiveness was very similar for either priming vaccine.

### Interpretation

These findings suggest that the booster offers very high levels of protection against symptomatic disease, at least in the short-term. Given the recent deployment of the booster programme in the UK, further follow-up is needed to understand how protection changes over time against both mild and severe disease. The slightly lower relative VE estimates of the booster in individuals with BNT162b2 as a primary course compared to the ChAdOx1-S in the primary analysis is likely due to the different baseline with higher VE after 2 doses of BNT162b2 as compared to ChAdOx1-S (8). When using unvaccinated controls, there was little difference in observed vaccine effectiveness of the booster dose with either primary course. We also observed a peak in testing at day 1 after the booster dose which is likely to be reactogenicity effects so shortly after the vaccine, as has been reported previously (12).

### Comparison with existing literature

In Israel a booster programme began in July 2021. Bar-On et al reported an adjusted rate ratios of 11.3 (10.4-12.3) against confirmed infection in booster dose recipients compared to those who received only 2 doses (equivalent to relative vaccine effectiveness of 91.2%) (13). This is slightly higher than the relative vaccine effectiveness that we report, which could reflect lower 2 dose vaccine effectiveness in the comparison group in Israel where a greater degree of waning has previously been reported. (9, 14, 15)Even greater protection has been reported in Israel against severe disease.(16)(13)

### Limitations

This is an observational study with a number of possible biases and should be interpreted with caution. The imperfect sensitivity PCR testing could cause misclassification of both cases and controls, which could attenuate vaccine effectiveness estimates. Many individuals will also have been previously infected so the VE measured is in the context of a population where many have already had natural exposure. We adjust for measured confounders, however, there may be residual confounding that we could not account for. Nevertheless, the similarity of the VE estimates using those with two doses and no booster as the baseline and using the 2-6 day period post booster as the baseline suggests that residual confounding is small. Use of the unvaccinated as a comparator to obtain absolute effectiveness is most susceptible to residual confounding as the totally unvaccinated population may differ in many ways to those who have had vaccine doses, many of which may lead to underestimation of VE (8). Despite this potential underestimation, using the unvaccinated comparator the absolute VE estimates were over 93%. Due to small numbers at this early stage of the booster roll out this study only assesses symptomatic disease, there is currently insufficient follow-up to estimate the effects on severe disease which leads to hospitalisation and death. For the same reason we are only able to report the early effects of the booster programme and it is not yet clear how long protection against COVID-19 following booster vaccination will last.

In these analyses, we were unable to report on the half dose (50µg) of mRNA-1273 vaccine due to low numbers as the majority of booster doses given in this period were BNT162b2. We were unable to assess the VE in all those targeted for a booster dose such as individuals with underlying health conditions, adult carers and adult household contacts of immunosuppressed individuals due to small numbers and difficultly identifying these individuals with the dataset.

#### Conclusions

Our study provides real world evidence of significant increased protection from the booster dose against symptomatic disease in those aged over 50 year of age irrespective of which primary course was received. This indicates that a high level of protection is achieved among older adults who are more vulnerable to severe COVID-19. This will be important in the 2021 to 2022 winter period when COVID-19 is likely to co-circulate alongside other respiratory viruses, including seasonal influenza virus.

## Data Availability

Data cannot be made publicly available
for ethical and legal reasons, i.e. public availability would compromise patient
confidentiality as data tables list single counts of individuals rather than aggregated
data

## Contributors

JLB, NA, and MR designed the study and developed the protocol and analysis plan. NA, FK and JS cleaned and analysed the data. JS drafted the manuscript. All authors contributed to the study design and revised the manuscript. The corresponding author attests that all listed authors meet authorship criteria and that no others meeting the criteria have been omitted. JLB and MR are the guarantors.

## Ethical approval

Surveillance of covid-19 testing and vaccination is undertaken under Regulation 3 of The Health Service (Control of Patient Information) Regulations 2002 to collect confidential patient information (www.legislation.gov.uk/uksi/2002/1438/regulation/3/ made) under Sections 3(i) (a) to (c), 3(i)(d) (i) and (ii) and 3(3). The study protocol was subject to an internal review by the Public Health England Research Ethics and Governance Group and was found to be fully compliant with all regulatory requirements. As no regulatory issues were identified, and ethical review is not a requirement for this type of work, it was decided that a full ethical review would not be necessary.

## Data sharing

No additional data available.

